# DERIVATION AND VALIDATION OF A CLINICAL SCORE TO PREDICT DEATH AMONG NON-PALLIATIVE COVID-19 PATIENTS PRESENTING TO EMERGENCY DEPARTMENTS: THE CCEDRRN COVID MORTALITY SCORE

**DOI:** 10.1101/2021.07.28.21261283

**Authors:** Corinne M. Hohl, Rhonda J. Rosychuk, Patrick M. Archambault, Fiona O’Sullivan, Murdoch Leeies, Éric Mercier, Gregory Clark, Grant D. Innes, Steven C. Brooks, Jake Hayward, Vi Ho, Tomislav Jelic, Michelle Welsford, Marco L.A. Sivilotti, Laurie J. Morrison, Jeffrey J. Perry, on behalf of the CCEDRRN investigators for the Network of Canadian Emergency Researchers and the Canadian Critical Care Trials Group

**Author notes:** (co-senior authors) (Laurie J. Morrison, Jeffrey J. Perry). **Corresponding Author:** Corinne M. Hohl, MD MHSc. **Funding Acknowledgement:** The network is funded by the Canadian Institutes of Health Research (447679), BC Academic Health Science Network Society, BioTalent Canada, Genome BC (COV024; VAC007), Ontario Ministry of Colleges and Universities (C-655-2129), the Saskatchewan Health Research Foundation (5357) and the Fondation CHU de Québec (Octroi #4007).

## Abstract

**Background:** Predicting mortality from coronavirus disease 2019 (COVID-19) using information available when patients present to the Emergency Department (ED) can inform goals-of-care decisions and assist with ethical allocation of critical care resources.

**Methods:** We conducted an observational study to develop and validate a clinical score to predict ED and in-hospital mortality among consecutive non-palliative COVID-19 patients. We recruited from 44 hospitals participating in the Canadian COVID-19 ED Rapid Response Network (CCEDRRN) between March 1, 2020 and January 31, 2021. We randomly assigned hospitals to derivation or validation, and pre-specified clinical variables as candidate predictors. We used logistic regression to develop the score in a derivation cohort, and examined its performance in predicting ED and in-hospital mortality in a validation cohort.

**Results:** Of 8,761 eligible patients, 618 (7·01%) died. The score included age, sex, type of residence, arrival mode, chest pain, severe liver disease, respiratory rate, and level of respiratory support. The area under the curve was 0·92 (95% confidence intervals [CI] 0·91–0·93) in derivation and 0·92 (95%CI 0·89–0·93) in validation. The score had excellent calibration. Above a score of 15, the observed mortality was 81·0% (81/100) with a specificity of 98·8% (95%CI 99·5–99·9%).

**Interpretation:** The CCEDRRN COVID Mortality Score is a simple score that accurately predicts mortality with variables that are available on patient arrival without the need for diagnostic tests.

**Trial registration:** Clinicaltrials.gov, NCT04702945

## INTRODUCTION

Throughout the coronavirus disease 2019 (COVID-19) pandemic, health systems around the world have been confronted with, and at times overwhelmed by high numbers of critically ill patients.(1,2) For every critically ill patient in the Emergency Department (ED), a number of less severely ill patients present for care, some of whom deteriorate later, placing additional pressure on resources. Accurate, disease-specific mortality prediction is needed to inform shared decision-making with patients and their families around the patients’ goals of care, and can allow healthcare systems to allocate resources in the most transparent, objective, and fair manner possible to save as many lives as possible, and facilitate timely access to palliative care, if needed.(3–5)

Numerous models have been developed to predict mortality from COVID-19, but most were at high-risk of bias.(6–8) Many were developed in small or non-representative patient samples, and enrolled patients from the early pandemic before evidence-based treatments had been identified, included palliative patients, censored outcomes, and had moderate predictive performance.(6) The ISARIC 4C Mortality score is the strongest of those developed.(9) However, it included palliative patients, limiting its utility in risk-stratifying non-palliative COVID-19 patients. In addition, the rule was developed using data from the early pandemic. Most other published rules use imaging or laboratory tests, which precludes their use as a first-line triage tool in the ED where decisions on the appropriateness of intubation and mechanical ventilation may have to be made on arrival.(9–14) Our objective was to develop and validate a clinical score that accurately predicts mortality among non-palliative COVID-19 patients, using clinical variables that are readily available on ED arrival.

## METHODS

### Study design and setting

The Canadian COVID-19 ED Rapid Response Network (CCEDRRN, pronounced “SEDrin”) is an ongoing multicentre pan-Canadian registry that enrolls consecutive eligible COVID-19 patients presenting to EDs in hospitals located in eight Canadian provinces, including the four most populous.(15) This study was approved by the research ethics boards of all participating institutions with a waiver of informed consent for enrolment. Model development and reporting followed TRIPOD standards.(16) Funders had no role in the collection, analysis, or interpretation of the data, the writing of the manuscript, or the decision to submit for publication. All authors vouch for the accuracy and completeness of the data, and for adherence to the protocol. CCEDRRN’s patient engagement committee reviewed and provided input into the development of the research question, the choice of outcomes and the study protocol, and reviewed the submitted manuscript. Patient partners were involved in developing CCEDRRN’s website and knowledge translation tools to disseminate study results.

### Study patients

Participating sites needed to demonstrate ≥99% compliance in enrolling consecutive eligible patients for their data to be included in this study. We included data from 44 of 50 CCEDRRN sites that met this criterion by the time of the data cut (Appendix Table 1).

**Table 1.**
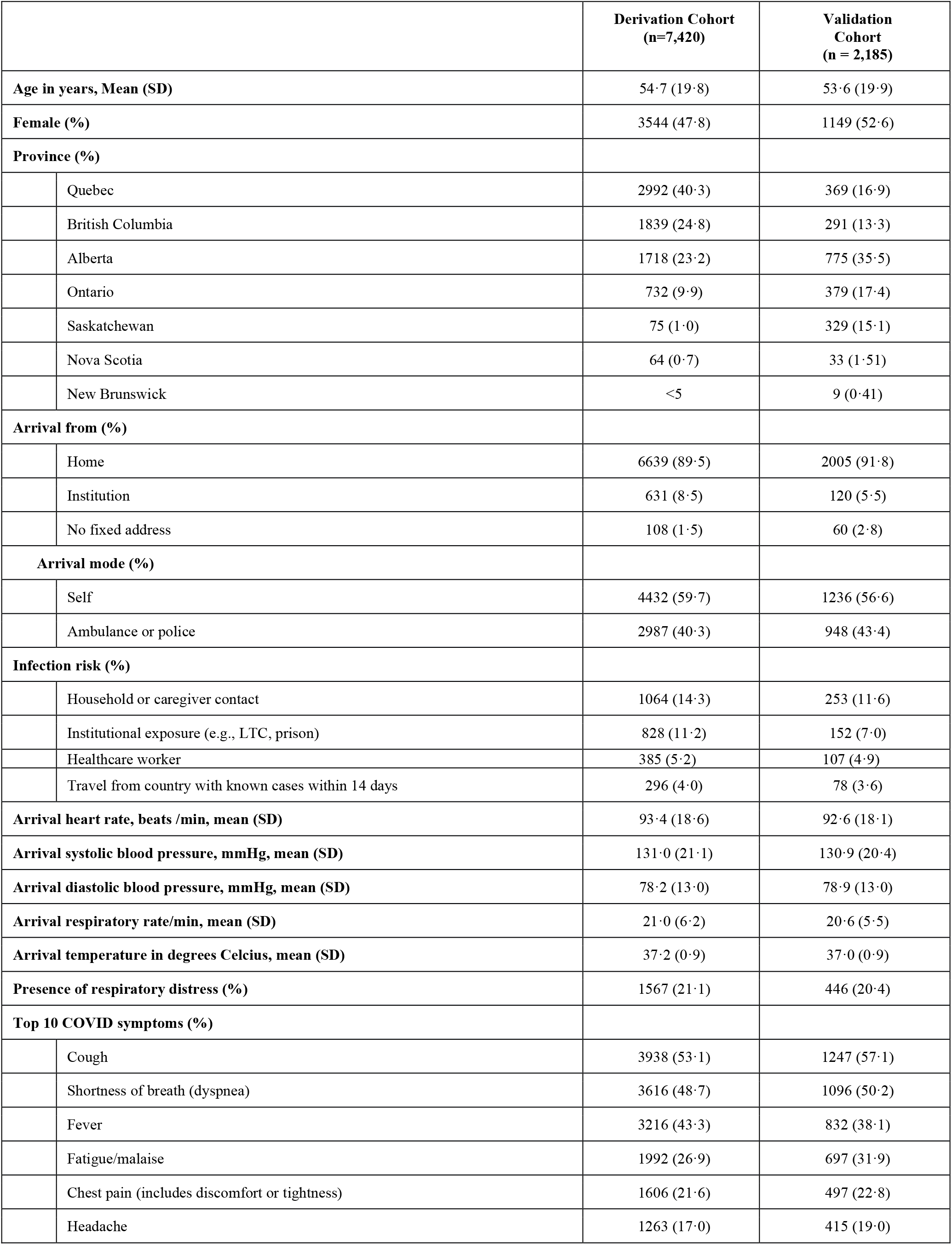
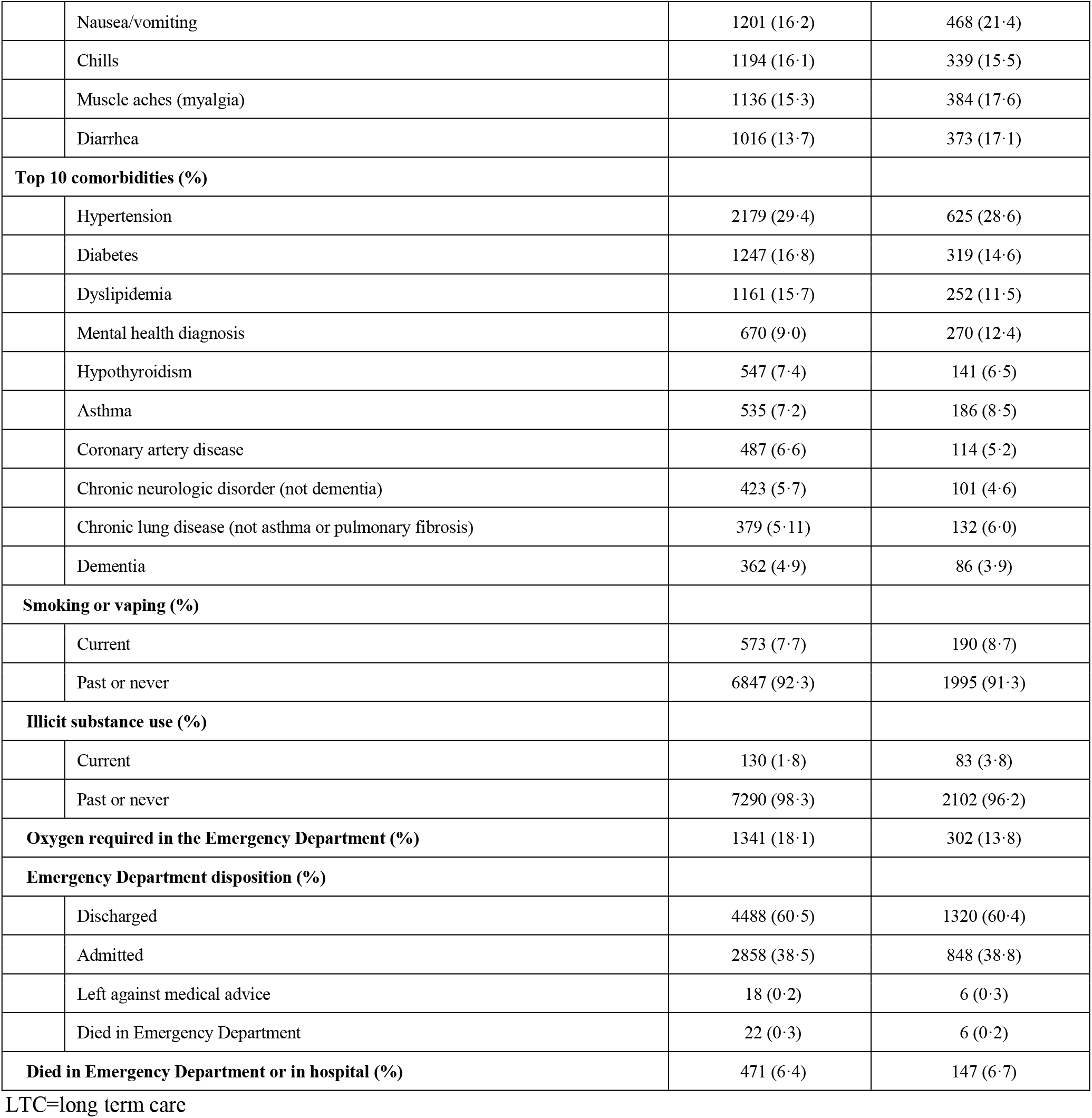
Characteristics and outcomes of patients in the derivation and validation cohorts.

|  |  | **Derivation Cohort**<br>**(n=7,420)** | **Validation Cohort**<br>**(n = 2,185)** |
| --- | --- | --- | --- |
| **Age in years, Mean (SD)** |  | 54·7 (19·8) | 53·6 (19·9) |
| **Female (%)** |  | 3544 (47·8) | 1149 (52·6) |
| **Province (%)** |  |  |  |
|  | Quebec | 2992 (40·3) | 369 (16·9) |
|  | British Columbia | 1839 (24·8) | 291 (13·3) |
|  | Alberta | 1718 (23·2) | 775 (35·5) |
|  | Ontario | 732 (9·9) | 379 (17·4) |
|  | Saskatchewan | 75 (1·0) | 329 (15·1) |
|  | Nova Scotia | 64 (0·7) | 33 (1·51) |
|  | New Brunswick | <5 | 9 (0·41) |
| **Arrival from (%)** |  |  |  |
|  | Home | 6639 (89·5) | 2005 (91·8) |
|  | Institution | 631 (8·5) | 120 (5·5) |
|  | No fixed address | 108 (1·5) | 60 (2·8) |
| **Arrival mode (%)** |  |  |  |
|  | Self | 4432 (59·7) | 1236 (56·6) |
|  | Ambulance or police | 2987 (40·3) | 948 (43·4) |
| **Infection risk (%)** |  |  |  |
|  | Household or caregiver contact | 1064 (14·3) | 253 (11·6) |
|  | Institutional exposure (e.g., LTC, prison) | 828 (11·2) | 152 (7·0) |
|  | Healthcare worker | 385 (5·2) | 107 (4·9) |
|  | Travel from country with known cases within 14 days | 296 (4·0) | 78 (3·6) |
| **Arrival heart rate, beats /min, mean (SD)** |  | 93·4 (18·6) | 92·6 (18·1) |
| **Arrival systolic blood pressure, mmHg, mean (SD)** |  | 131·0 (21·1) | 130·9 (20·4) |
| **Arrival diastolic blood pressure, mmHg, mean (SD)** |  | 78·2 (13·0) | 78·9 (13·0) |
| **Arrival respiratory rate/min, mean (SD)** |  | 21·0 (6·2) | 20·6 (5·5) |
| **Arrival temperature in degrees Celcius, mean (SD)** |  | 37·2 (0·9) | 37·0 (0·9) |
| **Presence of respiratory distress (%)** |  | 1567 (21·1) | 446 (20·4) |
| **Top 10 COVID symptoms (%)** |  |  |  |
|  | Cough | 3938 (53·1) | 1247 (57·1) |
|  | Shortness of breath (dyspnea) | 3616 (48·7) | 1096 (50·2) |
|  | Fever | 3216 (43·3) | 832 (38·1) |
|  | Fatigue/malaise | 1992 (26·9) | 697 (31·9) |
|  | Chest pain (includes discomfort or tightness) | 1606 (21·6) | 497 (22·8) |
|  | Headache | 1263 (17·0) | 415 (19·0) |
|  | Nausea/vomiting | 1201 (16·2) | 468 (21·4) |
|  | Chills | 1194 (16·1) | 339 (15·5) |
|  | Muscle aches (myalgia) | 1136 (15·3) | 384 (17·6) |
|  | Diarrhea | 1016 (13·7) | 373 (17·1) |
| **Top 10 comorbidities (%)** |  |  |  |
|  | Hypertension | 2179 (29·4) | 625 (28·6) |
|  | Diabetes | 1247 (16·8) | 319 (14·6) |
|  | Dyslipidemia | 1161 (15·7) | 252 (11·5) |
|  | Mental health diagnosis | 670 (9·0) | 270 (12·4) |
|  | Hypothyroidism | 547 (7·4) | 141 (6·5) |
|  | Asthma | 535 (7·2) | 186 (8·5) |
|  | Coronary artery disease | 487 (6·6) | 114 (5·2) |
|  | Chronic neurologic disorder (not dementia) | 423 (5·7) | 101 (4·6) |
|  | Chronic lung disease (not asthma or pulmonary fibrosis) | 379 (5·11) | 132 (6·0) |
|  | Dementia | 362 (4·9) | 86 (3·9) |
| **Smoking or vaping (%)** |  |  |  |
|  | Current | 573 (7·7) | 190 (8·7) |
|  | Past or never | 6847 (92·3) | 1995 (91·3) |
| **Illicit substance use (%)** |  |  |  |
|  | Current | 130 (1·8) | 83 (3·8) |
|  | Past or never | 7290 (98·3) | 2102 (96·2) |
| **Oxygen required in the Emergency Department (%)** |  | 1341 (18·1) | 302 (13·8) |
| **Emergency Department disposition (%)** |  |  |  |
|  | Discharged | 4488 (60·5) | 1320 (60·4) |
|  | Admitted | 2858 (38·5) | 848 (38·8) |
|  | Left against medical advice | 18 (0·2) | 6 (0·3) |
|  | Died in Emergency Department | 22 (0·3) | 6 (0·2) |
| **Died in Emergency Department or in hospital (%)** |  | 471 (6·4) | 147 (6·7) |

LTC=long term care

We included patients with confirmed COVID-19 who presented to the ED of a participating site between March 1, 2020 and January 31, 2021. We defined confirmed COVID-19 as patients presenting with ongoing COVID-19 symptoms and a positive nucleic acid amplification test (NAAT) for severe acute respiratory syndrome coronavirus-2 (SARS-CoV-2) obtained within 14 days prior to, or after their arrival in the ED. This allowed us to capture patients who were diagnosed in the community and subsequently presented to the ED, and those with early false negative tests that became positive. We also included patients presenting with COVID-19 symptoms and diagnosed with “confirmed COVID-19” to capture patients who were transferred into a CCEDRRN hospital whose NAAT at the sending site could not be confirmed, and patients who were presumed by treating clinicians to have COVID-19 despite persistently negative NAATs.

We excluded patients under 18 years of age, those whose goals of care precluded invasive mechanical ventilation, and patients transferred to a hospital outside of CCEDRRN, as we would have been unable to ascertain their outcomes (Figure 1). We followed patients for 30 days if they were discharged from the ED, or until hospital discharge if their admission lasted longer than 30 days.

**Figure 1.**
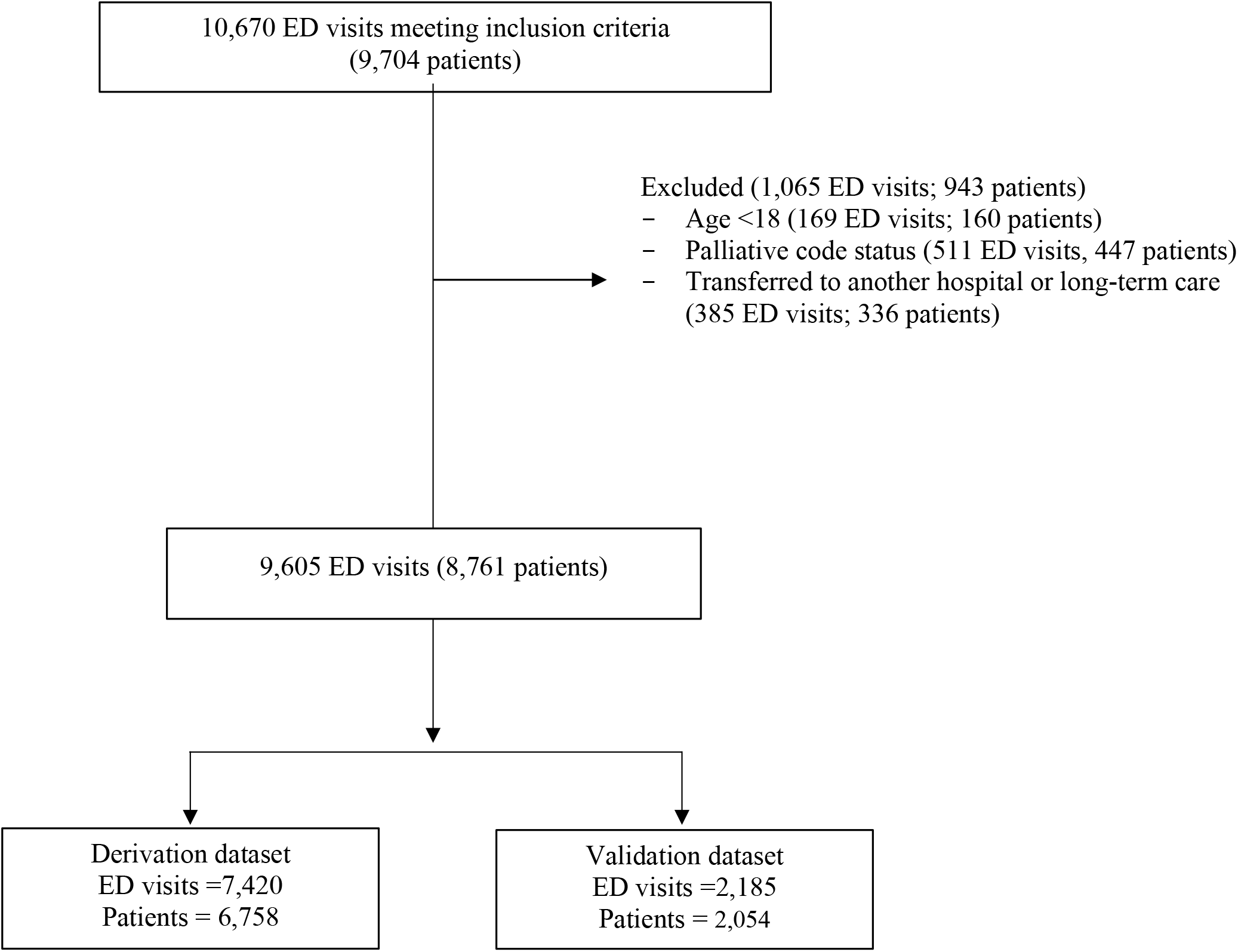
Flow diagram of included and excluded Emergency Department visits.

### Data collection

Trained research assistants abstracted data from electronic and paper-based medical records into a central, web-based REDCap database (Vanderbilt University; Nashville, TN, USA), and captured demographics, vital signs, symptoms, and comorbidities, COVID-19 exposure risk, diagnostic test results, and patient outcomes. We evaluated the inter-rater agreement of key predictor variables by comparing data collected retrospectively with prospective data.(15) The clinical prediction score was developed after all chart abstractions were complete; research assistants were thus unaware of which clinical variables would be candidate predictor variables.

### Outcome

The primary outcome was all-cause ED and in-hospital mortality. All patients had complete follow-up data at the time of the data cut. We categorized patients who were discharged from hospital as alive according to their latest hospitalization.

### Statistical analysis

#### Predictor variables

All candidate predictor variables were recorded in the ED record. We chose candidate predictors based on literature review and clinical knowledge. They included age, sex, pregnancy, type of residence, ED arrival mode, comorbidities, symptoms, arrival heart rate, systolic blood pressure, oxygen saturation, respiratory rate and Glasgow Coma Scale, ED oxygen delivery, lowest oxygen saturation, physician or nurse impression of respiratory distress, and use of alcohol, tobacco, vaping, and illicit substances (Appendix Table 2).

**Table 2.**
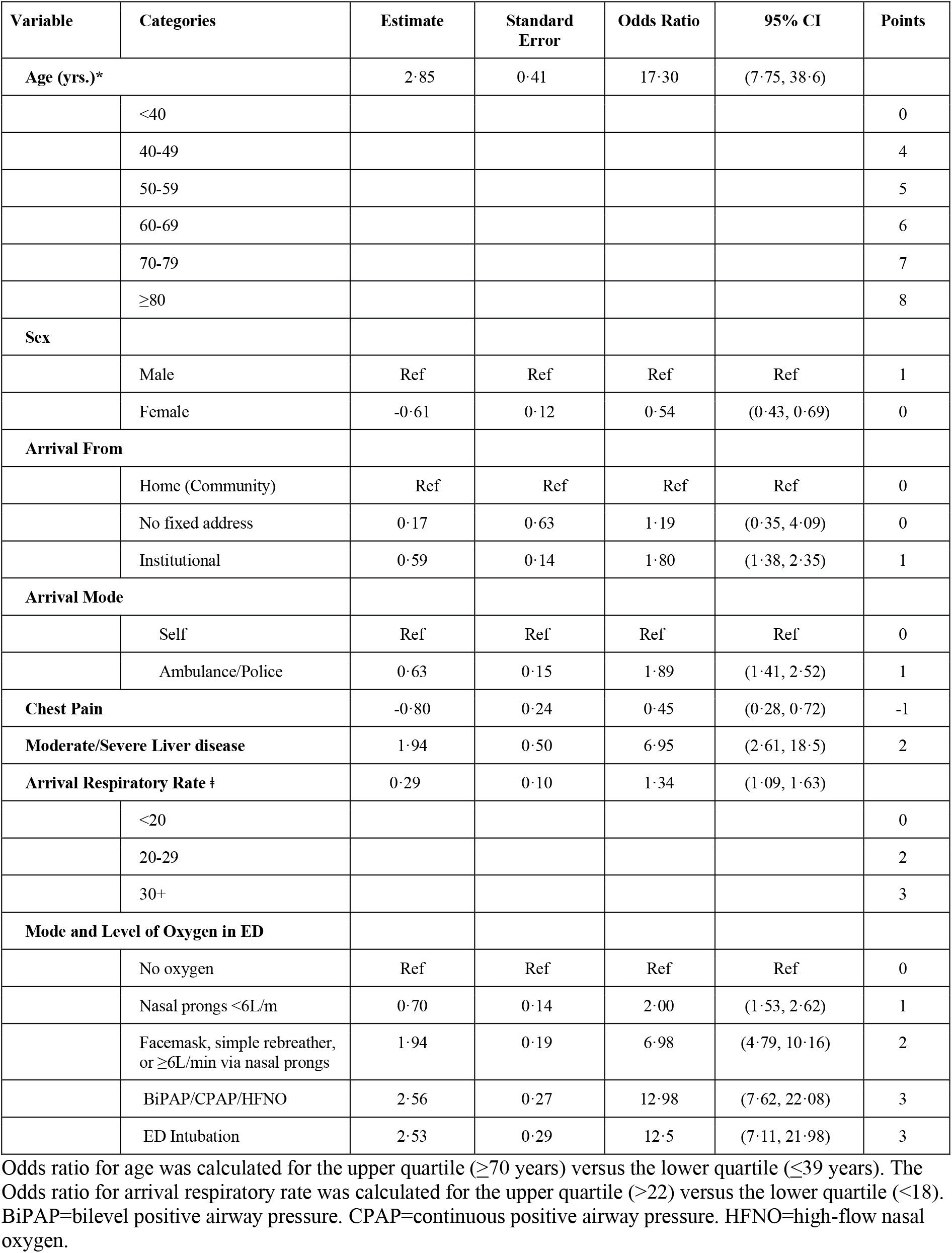
Adjusted associations between predictor variables and mortality, and points for the CCEDRRN COVID Mortality Score.

#### Model development and validation

We randomly assigned participating sites to derivation or validation, with the goal of assigning 75% of eligible patients and outcome events to derivation, and the remaining to validation. We examined candidate predictors for co-linearity, and missing and extreme values in the derivation cohort. A few variables had missing values (systolic blood pressure had the most missing at 4.7%; Appendix Table 2). We used five multiple imputations for predictors if missing categorical data could not reasonably be assumed to be absent (e.g., missing documentation of illicit substance use was classified as no substance use). The initial logistic regression model considered all candidate predictors, with continuous predictors fit with restricted cubic splines with three knots. We assessed the strengths of associations between predictors and the outcome using an analysis of variance (ANOVA) plot to inform the degrees of freedom to allocate to each predictor. No additional knots were allocated to continuous predictors. We used a fast step-down procedure to reduce the model to key predictors. We conducted an internal bootstrap validation with 1,000 bootstrap samples to provide an optimism-corrected area under the receiver-operating characteristic (AUC). We categorized continuous predictors based on the relationship between the spline function and outcome. To enable easy clinical use, we categorized age into decades, and arrival respiratory rate cut points of 20 and 30, and assigned integer points to be added to calculate the score. We used a nomogram to assign points to form a score that ranged from −1 to 17. We calculated the sensitivity and specificity at different point thresholds, along with the score’s discrimination and calibration. We validated the model in a cohort of geographically distinct sites that were not part of derivation, and used a single imputation for the few missing respiratory rates (4% were missing). We assessed outcomes independently for ED visits, irrespective of potential subsequent visits leading to death.

#### Validation of previously published models

We used our study cohort to externally validate other risk prediction tools: the SEIMC score,^12^ the 4C Mortality Score,^10^ and the VACO Index.^20^ We chose these three because they performed well in validation, and the majority of their predictors were available in our data. We calculated the AUCs for these risk prediction tools using cases with complete data on as many predictors as possible.(17)

We performed analyses in R using the rms package and used the pmsampsize package for sample size determination.(18) To ensure patient privacy, a cell size restriction policy prohibited reporting counts of less than five.

#### Sample size

Assuming an event rate of less than 10%, shrinkage of 0.9, and a conservative Cox-Snell R-squared of 0.1, 8.5 events per degree of freedom were required for reliable prediction modeling in the derivation cohort.(19) The 42 candidate predictor variables had 49 degrees of freedom indicating 417 events were required. In the derivation cohort, there were 6,758 patients who made 7,420 ED visits. The derivation cohort exceeded the number of events required.

## RESULTS

We assessed 9,704 consecutive COVID-19 patients who made 10,670 ED visits between March 1, 2020 and January 31, 2021 (Figure 1). We excluded 943 patients who met one or more exclusion criteria, and included 8,761 patients who made 9,605 visits in our analyses. The follow-up time was 30 days for discharged patients and between 30 and 229 days for admitted patients. Of these, 618 (7.0%) died in ED or hospital and met the primary outcome. In the derivation cohort, 6,758 patients made 7,420 ED visits to 32 sites (Appendix Table 1). In the validation cohort, 2,054 patients made 2,185 ED visits to 14 different sites. In the derivation cohort 2,705 (36.5%) patients presented during the early pandemic, between March 1 and June 30, 2020, and 4,711 (63.7%) between July 1, 2020 and January 31, 2021 (Table 1). In the validation cohort, 6158 (28.4%) presented during the early pandemic, and 1,567 (71.7%) after July 1, 2020.

In derivation, the step-down procedure produced a final model with eight variables (Table 2). The derived model had an optimism-corrected AUC of 0.92. The resulting risk score ranged from −1 to 17. The derivation cohort was well distributed across the CCEDRRN COVID Mortality Score range, and had excellent calibration (calibration intercept of 0 and slope of 1) and discrimination with an AUC of 0.92 (95% CI 0.90 to 0.92, Appendix Figure 1).

The CCEDRRN COVID Mortality Score had similar performance in validation. The validation cohort was also distributed across the score ranges, had excellent calibration (calibration intercept of 0 and slope of 1) and discrimination (AUC of 0.92 [95% CI 0.89 to 0.93], Figure 2).

**Figure 2.**
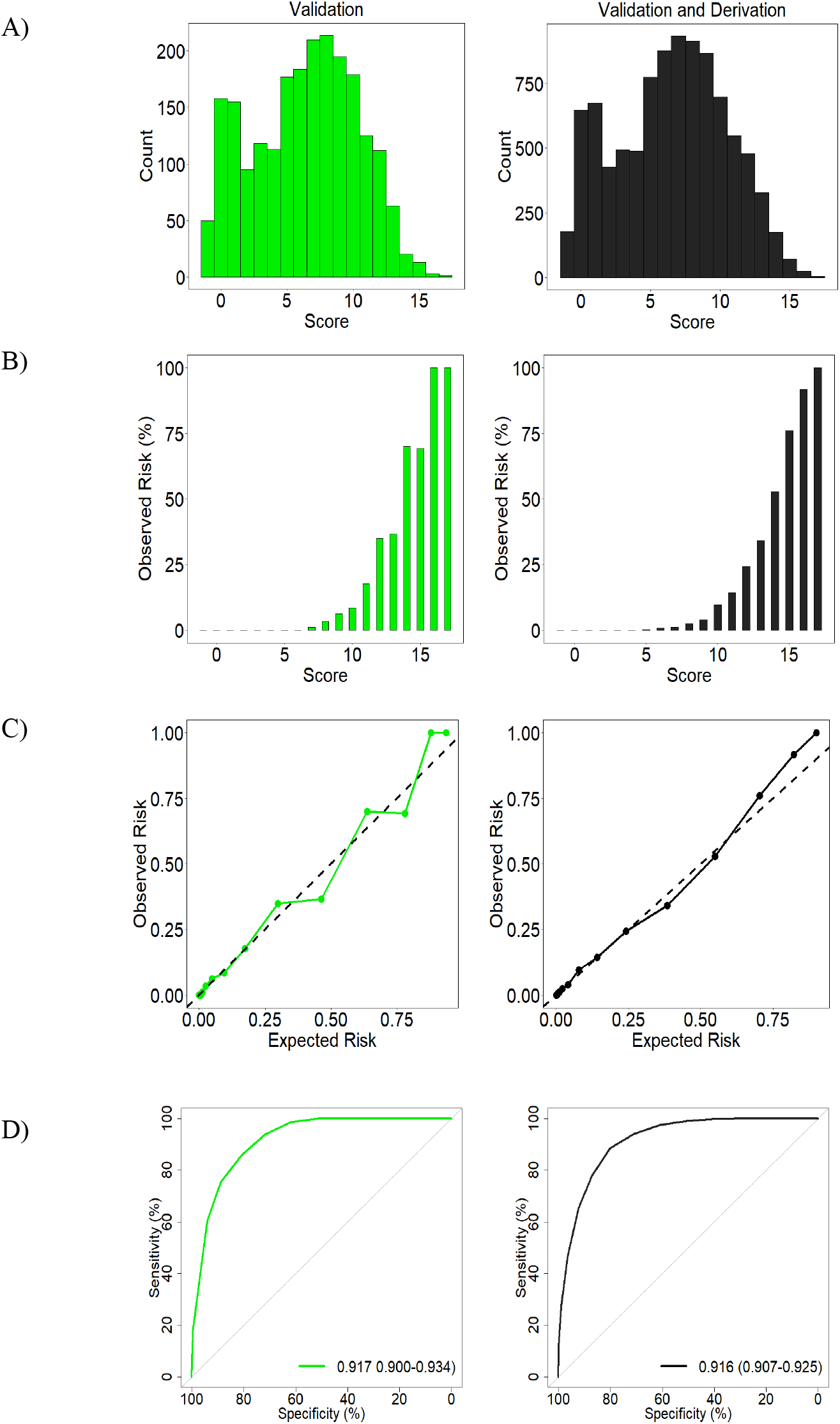
Distribution and performance of the CCEDRRN COVID Mortality Score in the validation cohort (left panel) and combined derivation and validation cohorts (right panel): A) distribution of the score, B) observed in-hospital mortality across the range of the score, C) predicted versus observed probability of in-hospital mortality, and D) receiver operating characteristic curve with area under the curve (AUC) and associated 95% confidence interval.

The score had excellent performance across a range of thresholds to rule in and rule out in-hospital mortality (Appendix Tables 4 and 5). These results suggest that scores less than or equal to six would categorize patients at low risk of in-hospital mortality, with a negative predicted value of 99.9%. Patients in the low-risk group had an in-hospital mortality of 0.1%. For scores greater than or equal to 15, the observed in-hospital mortality was 81.0% and the CCEDRRN COVID Mortality Score would categorize patients at high risk of in-hospital mortality, with a specificity of 99.8% and positive predictive value of 81.0%.

We conducted an external validation of three risk scores (Figure 3, Appendix Table 5). These scores performed well for the patients with data available yielding AUCs of nearly 0.88, below the AUC of the CCEDRRN COVID Mortality Score (unadjusted for multiple testing and using the validation cohort: AUC for the CCEDRRN COVID Mortality Score was higher compared to the SEIMC score [p=0.035] and the VACO Index [p=0.002], with no evidence of a difference for the 4C Mortality Score [p=0.072]).

**Figure 3.**
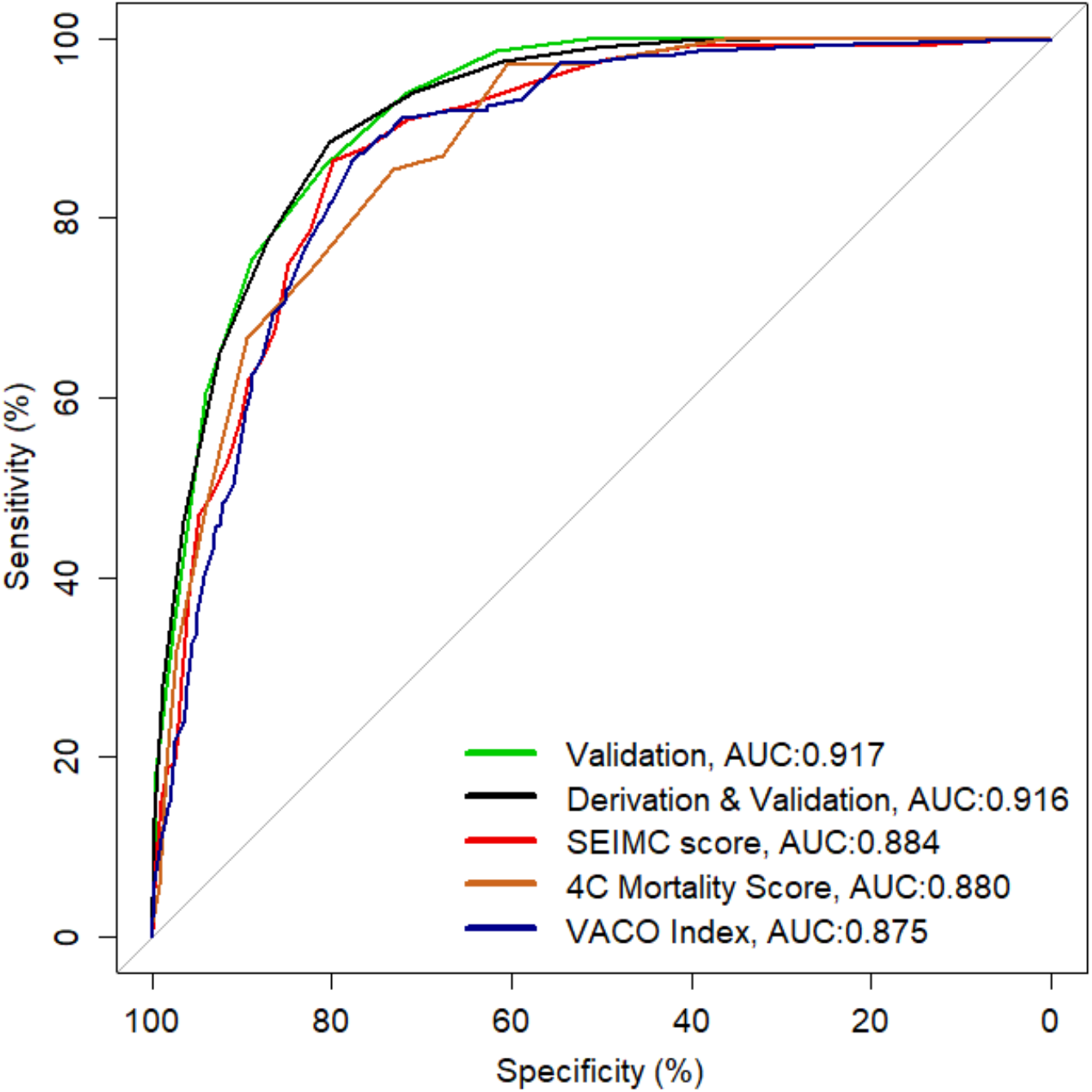
Receiver operator curves and area under the curve (AUC) for the CCEDRRN COVID Mortality Score in the derivation and validation cohorts, the SEIMC score (Berenguer et al.),(11) the 4C Mortality Score (Knight et al.),(9) and the VACO Index (King et al.) in the validation cohort.(20)

## INTERPRETATION

### Main results

We derived and validated a parsimonious and simple score to predict in-hospital mortality among non-palliative patients presenting to EDs with COVID-19: the CCEDRRN COVID Mortality Score. We found that eight readily available clinical variables that can be ascertained at the bedside shortly after ED arrival to accurately predict mortality. The CCEDRRN COVID Mortality Score had excellent calibration and discrimination in a geographically distinct cohort of patients who presented to other sites. The CCEDRRN COVID Mortality Score can be used as a highly sensitive score to rule out in-hospital mortality in low-risk patients with a score up to and including six. It can also be used as a rule-in score with scores of 15 or higher being highly specific for in-hospital mortality.

Critically ill COVID-19 patients typically require aggressive medical management in the ED shortly after their arrival. Being able to accurately and reliably predict mortality risk on arrival before endotracheal intubation occurs can offer the opportunity to inform discussions about patients’ goals of care, and facilitate early high-quality end of life care for patients most likely to die despite maximum medical intervention. Accurate mortality prediction may be essential when surging cases threaten to overwhelm critical care resources. In those rare situations, the CCEDRRN COVID Mortality Score can guide allocation of scarce resources.

### Explanation of the findings

Our model has strengths compared to prior models.(6) We developed the CCEDRRN COVID Mortality Score using simple and readily available variables at the bedside. As a result, our model could be externally validated for use in remote areas without access to laboratory testing or imaging, and in low-income countries where access may be limited or costly. In contrast to prior models, we excluded patients with palliative goals of care, for whom invasive mechanical ventilation was not offered to ensure our model did not predict patients who were expected to succumb, or ineligible for the highest level of critical care.(9–11,13,14,20–24) This avoids the potential for self-fulfilling prophecy bias, whereby the prognostic model predicts the outcome that occurred as a result of a decision to withhold life-sustaining measures.(25–27) Prior models were derived or validated during the early pandemic while COVID-19 testing was restricted to those with severe disease, and did not include consecutive eligible patients, both of which may have resulted in selection bias. Mortality in prior studies ranged from 13% to 30%,(9,11,12,14,22–24) in contrast to 7% in our study.

Our rule’s predictive ability depends mostly on age, the patient’s respiratory status. The only comorbidity retained in the final model was moderate to severe liver disease. Two other rules also identified liver disease as a mortality risk factor, perhaps due to the potential for virus-induced liver inflammation.(14,20,28) Other prognostic decision rules have used similar analytic approaches,(20,22,23) but had lower predictive performance with c-statistics ranging from 0.80 to 0.82, and were based on patients from the early pandemic. Other rules have incorporated measures of hypoxemia or respiratory support, corroborating their strong predictive power.(9– 13,20–24) Goodacre et al. identified performance status as a risk factor for increased mortality. In our dataset, this was most closely reflected by arrival from long-term care, which reflected patients with lower performance status.(23)

### Limitations of the study

Canada represents a culturally diverse country that offers its citizens universal health coverage. Our model needs to be externally validated in other health systems. Our model predicts in-hospital mortality, and may have missed out-of-hospital deaths that occurred after discharge; however, these are believed to be rare. Our model was based on patients with confirmed COVID-19. While many patients presented to the ED with NAAT confirmed

COVID-19, validation in a cohort of patients with suspected COVID-19 is needed.

### Future directions in the area of study

As vaccination campaigns roll out around the world, the performance of risk tools will need to be evaluated in patients who are vaccinated and may have a different risk of dying from COVID-19.

### Conclusion

In summary, the CCEDRRN COVID Mortality Score is a simple clinical risk score that can be applied in the ED at the bedside to predict a patient’s mortality risk. This tool can be used to inform goals of care decisions.

## Data Availability

The CCEDRRN network endorses the guidance put forth by the World Health Organization to enable data sharing to optimize learning. CCEDRRN accepts applications for access to data by external investigators, prioritizing data requests by network Members.

## Authors’ contributions

CMH, RJR, JJP, LJM, PA and SCB conceived the study, with input on the design and selection of variables from all other contributors. CMH, LJM, PA, and SCB obtained funding on behalf of the CCEDRRN investigators. CMH, PA, SCB, EM, MW, JH, TJ, VH, and GC managed data collection along with other members of the CCEDRRN, and verify the accuracy of underlying data. CMH, RJR, FOS and JJP developed the analytic plan. FOS performed the analysis, with assistance from RJR, CMH and JJP, including accessing and verification of underlying data. All contributors provided input on the interpretation our findings. CMH and PA drafted the manuscript. All authors reviewed and provided critical input to the final manuscript.

## Acknowledgements

We gratefully acknowledge the assistance of Ms. Amber Cragg in the preparation of this manuscript. We thank the UBC clinical coordinating centre staff, the UBC legal, ethics, privacy and contract staff and the research staff at each of the participating institutions in the network outlined in the attached Supplement. The network would not exist today without the dedication of these professionals.

Thank you to all of our patient partners who shared their lived experiences and perspectives to ensure that the knowledge we co-create addresses the concerns of patients and the public. Creating the largest network of collaboration across Canadian Emergency Departments would not have been feasible without the tireless efforts of Emergency Department Chiefs, and research coordinators and research assistants at participating sites.

Finally, our most humble and sincere gratitude to all of our colleagues in medicine, nursing, and the allied health professions who have been on the front lines of this pandemic from day one staffing our ambulances, Emergency Departments, ICUs and hospitals bravely facing the risks of COVID-19 to look after our fellow citizens and after one another. We dedicate this network to you.

## APPENDICES

**Appendix Table 1.** Characteristics and enrolment periods for participating sites.

| Hospital | Province | Start Date | End Date | Hospital Type |  | Derivation cohort | Validation cohort |
| --- | --- | --- | --- | --- | --- | --- | --- |
|  |  |  |  | Rural/<br>Urban | Teaching | n | n |
| Vancouver General Hospital | BC | Mar 1, 2020 | Jun 30, 2020 | Urban | Teaching | 91 |  |
| Lions Gate Hospital | BC | Mar 1, 2020 | Sep 19, 2020 | Urban | NT |  | 142 |
| Saint Paul's Hospital | BC | Mar 1, 2020 | Mar 20, 2020 | Urban | Teaching |  | 59 |
| Mount St Joseph's Hospital | BC | Mar 1, 2020 | Oct 31, 2020 | Urban | NT |  | 90 |
| Surrey Memorial Hospital | BC | Mar 19, 2020 | Sep 20, 2020 | Urban | Teaching | 895 |  |
| Royal Columbian Hospital | BC | Mar 1, 2020 | Nov 15, 2020 | Urban | Teaching | 221 |  |
| Abbotsford Regional Hospital | BC | Apr 20, 2020 | Jul 15, 2020 | Urban | NT | 329 |  |
| Eagle Ridge Hospital | BC | Mar 1, 2020 | Nov 30, 2020 | Urban | NT | 162 |  |
| Royal Inland Hospital | BC | Mar 1, 2020 | Nov 30, 2020 | Urban | NT | 35 |  |
| Kelowna General Hospital | BC | Mar 1, 2020 | Nov 2, 2020 | Urban | Teaching | 105 |  |
| University of Alberta Hospital | AB | Apr 8, 2020 | Oct 5, 2020 | Urban | Teaching | 263 |  |
| Foothills Medical Centre | AB | Mar 1, 2020 | Oct 14, 2020 | Urban | Teaching | 511 |  |
| Rockyview General Hospital | AB | Mar 1, 2020 | Oct 24, 2020 | Urban | Teaching |  | 446 |
| Peter Lougheed Centre | AB | Mar 1, 2020 | Oct 17, 2020 | Urban | Teaching | 829 |  |
| South Health Campus | AB | Mar 1, 2020 | Oct 17, 2020 | Urban | Teaching |  | 329 |
| Royal Alexandra Hospital | AB | Mar 1, 2020 | May 7, 2020 | Urban | Teaching |  | 180 |
| Northeast Community Health Centre | AB | Mar 1, 2020 | Nov 06, 2020 | Urban | Teaching | 115 |  |
| St Paul's Hospital | SK | Mar 17, 2020 | Dec 5, 2020 | Urban | Teaching |  | 149 |
| Royal University Hospital | SK | Mar 17, 2020 | Sep 20, 2020 | Urban | Teaching | 33 |  |
| Saskatoon City Hospital | SK | Mar 17, 2020 | Nov 17, 2020 | Urban | Teaching | 42 |  |
| Sunnybrook Hospital | ON | May 14, 2020 | Jul 31, 2020 | Urban | Teaching |  | 86 |
| The Ottawa Hospital - Civic | ON | Mar 1, 2020 | Nov 30, 2020 | Urban | Teaching | 214 |  |
| The Ottawa Hospital - General | ON | Mar 1, 2020 | Oct 31, 2020 | Urban | Teaching |  | 293 |
| Kingston General Hospital | ON | Mar 1, 2020 | Dec 31, 2020 | Urban | Teaching | 32 |  |
| Hamilton General Hospital | ON | Mar 1, 2020 | Aug 31, 2020 | Urban | Teaching | 46 |  |
| Jurvinski Hospital | ON | Mar 1, 2020 | Aug 31, 2020 | Urban | Teaching | 42 |  |
| Health Science North | ON | May 14, 2020 | Jul 25, 2020 | Urban | NT | 10 |  |
| University Hospital - LHSC | ON | Mar 1, 2020 | Nov 30, 2020 | Urban | Teaching | 312 |  |
| Toronto Western Hospital | ON | Sep 1, 2020 | Sep 19, 2020 | Urban | Teaching | 76 |  |
| HôtelHôtel-Dieu de Lévis | QC | Mar 1, 2020 | Nov 17, 2020 | Urban | NT | 267 |  |
| Jewish General Hospital | QC | Mar 1, 2020 | May 3, 2020 | Urban | Teaching | 815 |  |
| CHUL | QC | Mar 1, 2020 | Nov 15, 2020 | Urban | Teaching |  | 57 |
| Royal Victoria Hospital | QC | Mar 1, 2020 | Oct 31, 2020 | Urban | Teaching | 789 |  |
| Hôpital de l'Enfant-Jésus | QC | Mar 1, 2020 | May 4, 2020 | Urban | Teaching | 51 |  |
| Hôpital du Saint-Sacrement | QC | Mar 1, 2020 | Nov 15, 2020 | Urban | Teaching | 33 |  |
| Hôpital Saint-François d'Assise | QC | Mar 1, 2020 | Nov 15, 2020 | Urban | Teaching | 56 |  |
| Hôtel-Dieu de Québec | QC | Mar 1, 2020 | Nov 15, 2020 | Urban | Teaching | 21 |  |
| IUCPQ | QC | Mar 1, 2020 | Nov 15, 2020 | Urban | Teaching |  | 312 |
| Hôpital du Sacré-Coeur | QC | Mar 17, 2020 | Jun 11, 2020 | Urban | Teaching | 664 |  |
| Montreale General Hospital | QC | Mar 1, 2020 | Nov 30, 2020 | Urban | Teaching | 296 |  |
| Saint John Regional Hospital | NB | Mar 1, 2020 | Nov 30, 2020 | Urban | Teaching |  | 9 |
| Halifax Infirmary | NS | Mar 1, 2020 | Nov 24, 2020 | Urban | Teaching | 37 |  |
| Dartmouth General Hospital | NS | Mar 1, 2020 | Nov 24, 2020 | Com | NT | 16 |  |
| Hants Community Hospital | NS | Mar 1, 2020 | Nov 24, 2020 | Rural | NT |  | <5 |
| Cobequid Community Health Centre | NS | Mar 1, 2020 | Nov 24, 2020 | Com | NT | 11 |  |
| Secondary Assessment Centers | NS | Mar 26, 2020 | May 15, 2020 | Urban | Teaching |  | 31 |

BC=British Columbia, AB=Alberta, SK=Saskatchewan, ON=Ontario, QC= Québec, NB:=New Brunswick, NS= Nova Scotia, LHSC=London Health Sciences Centre, CHUL=Centre Hospitalier de l'Université Laval, IUCPQ=Institut universitaire de cardiologie et de pneumologie de Québec, Com=community

**Appendix Table 2.** Candidate variables for entry into regression model.

| Variable | Definition | N (%)<br>Missing |
| --- | --- | --- |
| **Demographics** |  |  |
| Age | Age in years | 0 (0) |
| Sex | Male, Female, Other | 0 (0) |
| Arrival from |  | 0 (0) |
|  | Home + Inter-hospital transfer + other (not clearly documented) | 42 (0.6) |
|  | Single room + no fixed address + shelter |  |
|  | Institutional living: long-term care/rehab + correctional |  |
| **Emergency department variables** |  |  |
| ED arrival mode |  | 0 (0) |
| Ambulance: | arrived by ambulance |  |
| Self/police | self-transported or transported to ED by police |  |
| Arrival heart rate | beats/minute | 131 (1.8) |
| Arrival respiratory rate | breaths/minute | 324 (4.4) |
| Arrival oxygen saturation | % | 131 (1.8) |
| Arrival systolic blood pressure | mmHg | 352 (4.7) |
| Fever | Temperature $\geq 37.5$ OR self-reported fever | 0 (0) |
| Respiratory distress | Increased work of breathing documented by treating clinician and Patient-reported symptom of shortness of breath as documented by treating clinician | 0 (0) |
| Bloodwork in ED | Yes/No |  |
| Supplemental oxygen delivered in the ED | Yes/No | 0 (0) |
| **COVID symptoms** |  |  |
| Chest pain (includes discomfort or tightness) | Patient-reported symptom as documented by treating clinician | 0 (0) |
| Chills | Patient-reported symptom as documented by treating clinician | 0 (0) |
| Cough | Patient-reported symptom as documented by treating clinician | 0 (0) |
| Dysgeusia/anosmia | Patient-reported symptom as documented by treating clinician | 0 (0) |
| Fatigue/malaise | Patient-reported symptom as documented by treating clinician | 0 (0) |
| Headache | Patient-reported symptom as documented by treating clinician | 0 (0) |
| Hemoptysis (bloody sputum) | Patient-reported symptom as documented by treating clinician | 0 (0) |
| Muscle aches (myalgia) | Patient-reported symptom as documented by treating clinician | 0 (0) |
| No reported symptoms | Patient-reported symptom as documented by treating clinician | 0 (0) |
| Current tobacco user | Documented current tobacco use | 0 (0) |
| Current illicit user | Documented methamphetamine, opioid or other illicit drug use | 0 (0) |
| Pregnant | Documented current pregnancy | 0 (0) |
| Arrival confusion | Patient-reported symptom of altered consciousness/confusion or Glasgow Coma Score of $<14$ | 0 (0) |
| Covid Symptom-gastric | Patient-reported symptom of diarrhea and nausea/vomiting | 0 (0) |
| Mode of oxygen delivery combined with oxygen received in the ED | Documented mode of oxygen delivery, combined with maximum oxygen received if given nasal prongs. Most invasive method of oxygen delivery selected if more than one documented. | 0 (0) |
|  | Intubation |  |
|  | BiPap + CPAP + HFNO |  |
| | Facemask + simple rebreather + non-rebreather + nasal prongs with $> 6L/min$ | |
| | Nasal prongs $<5L/min$ | |
|  | No oxygen |  |
| **Common Comorbid Conditions, n (%)** |  |  |
| Active malignant neoplasm (cancer) | The diagnosis, conditions, problem or circumstances for the patient's ED visit that is in addition to the main diagnosis. | 0 (0) |
| Coronary artery disease | The diagnosis, conditions, problem or circumstances for the patient's ED visit that is in addition to the main diagnosis. | 0 (0) |
| Congestive heart failure | The diagnosis, conditions, problem or circumstances for the patient's ED visit that is in addition to the main diagnosis. | 0 (0) |
| Hypertension | The diagnosis, conditions, problem or circumstances for the patient's ED visit that is in addition to the main diagnosis. | 0 (0) |
| Asthma | The diagnosis, conditions, problem or circumstances for the patient's ED visit that is in addition to the main diagnosis. | 0 (0) |
| Chronic kidney disease | The diagnosis, conditions, problem or circumstances for the patient's ED visit that is in addition to the main diagnosis. | 0 (0) |
| Dialysis | The diagnosis, conditions, problem or circumstances for the patient's ED visit that is in addition to the main diagnosis | 0 (0) |
| Diabetes | The diagnosis, conditions, problem or circumstances for the patient's ED visit that is in addition to the main diagnosis | 0 (0) |
| Moderate/severe liver disease | The diagnosis, conditions, problem or circumstances for the patient's ED visit that is in addition to the main diagnosis | 0 (0) |
| Organ transplant | The diagnosis, conditions, problem or circumstances for the patient's ED visit that is in addition to the main diagnosis | 0 (0) |
| Dementia | The diagnosis, conditions, problem or circumstances for the patient's ED visit that is in addition to the main diagnosis | 0 (0) |
| Rheumatologic disorder | The diagnosis, conditions, problem or circumstances for the patient's ED visit that is in addition to the main diagnosis | 0 (0) |
| Obesity (clinical impression) | The diagnosis, conditions, problem or circumstances for the patient's ED visit that is in addition to the main diagnosis | 0 (0) |
| Atrial fibrillation | The diagnosis, conditions, problem or circumstances for the patient's ED visit that is in addition to the main diagnosis | 0 (0) |

**Appendix Table 3.**
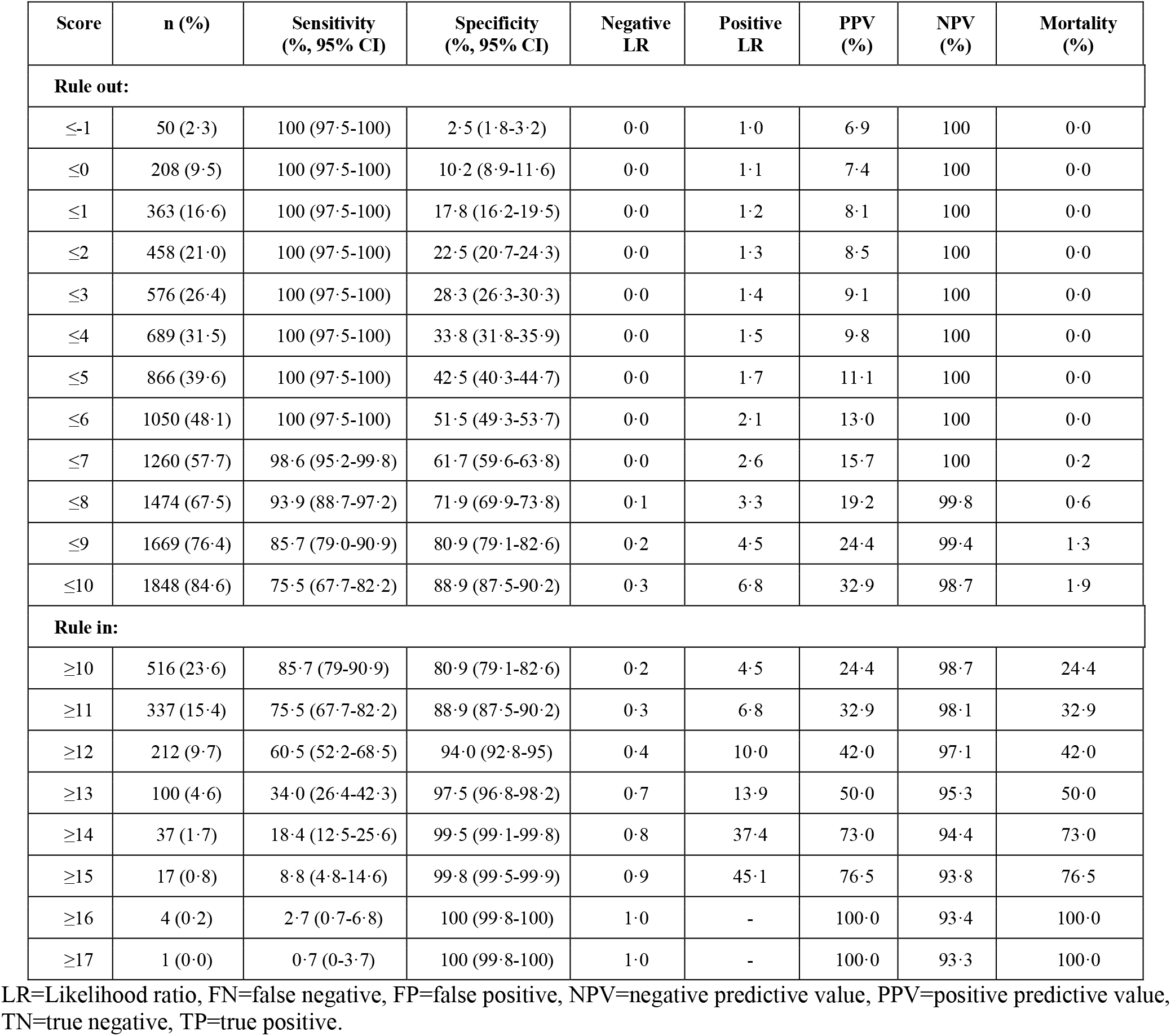
Performance of the CCEDRRN COVID Mortality Score to rule out and rule in in-hospital mortality at different cut-off values in the validation cohort.

**Appendix Table 4.** Performance of the CCEDRRN COVID Mortality Score to rule out and rule in in-hospital mortality at different cut-off values in the combined derivation and validation cohorts.

| Score | n (%) | Sensitivity<br>(%, 95% CI) | Specificity<br>(%, 95% CI) | Negative<br>LR | Positive<br>LR | PPV<br>(%) | NPV<br>(%) | Mortality<br>(%) |
| --- | --- | --- | --- | --- | --- | --- | --- | --- |
| **Rule out:** |  |  |  |  |  |  |  |  |
| ≤-1 | 178 (1·9) | 100·0 (99·4-100·0) | 2·0 (1·7-2·3) | 0·0 | 1·0 | 6·6 | 100 | 0·0 |
| ≤0 | 825 (8·6) | 100·0 (99·4-100·0) | 9·2 (8·6-9·8) | 0·0 | 1·1 | 7·0 | 100 | 0·0 |
| ≤1 | 1499 (15·6) | 100·0 (99·4-100·0) | 16·7 (15·9-17·5) | 0·0 | 1·2 | 7·6 | 100 | 0·0 |
| ≤2 | 1926 (20·1) | 100·0 (99·4-100·0) | 21·4 (20·6-22·3) | 0·0 | 1·3 | 8·0 | 100 | 0·0 |
| ≤3 | 2420 (25·2) | 100·0 (99·4-100·0) | 26·9 (26·0-27·9) | 0·0 | 1·4 | 8·6 | 100 | 0·0 |
| ≤4 | 2910 (30·3) | 100·0 (99·4-100·0) | 32·4 (31·4-33·4) | 0·0 | 1·5 | 9·2 | 100 | 0·0 |
| ≤5 | 3685 (38·4) | 99·8 (99·1-100·0) | 41·0 (40·0-42·0) | 0·0 | 1·7 | 10·4 | 100 | 0·0 |
| ≤6 | 4563 (47·5) | 99·0 (97·9-99·6) | 50·7 (49·7-51·7) | 0·0 | 2·0 | 12·1 | 99·9 | 0·1 |
| ≤7 | 5497 (57·2) | 97·4 (95·8-98·5) | 61·0 (60·0-62·0) | 0·0 | 2·5 | 14·7 | 99·7 | 0·3 |
| ≤8 | 6410 (66·7) | 94·0 (91·8-95·7) | 70·9 (70·0-71·9) | 0·1 | 3·2 | 18·2 | 99·4 | 0·6 |
| ≤9 | 7276 (75·8) | 88·5 (85·7-90·9) | 80·2 (79·3-81·0) | 0·1 | 4·5 | 23·5 | 99·0 | 1·0 |
| ≤10 | 7974 (83·0) | 77·7 (74·2-80·9) | 87·2 (86·5-87·9) | 0·3 | 6·1 | 29·4 | 98·3 | 1·7 |
| **Rule in:** |  |  |  |  |  |  |  |  |
| ≥10 | 2329 (24·2) | 88·5 (85·7-90·9) | 80·2 (79·3-81·0) | 0·1 | 4·5 | 23·5 | 99·0 | 23·5 |
| ≥11 | 1631 (17·0) | 77·7 (74·2-80·9) | 87·2 (86·5-87·5) | 0·3 | 6·1 | 29·4 | 98·3 | 29·4 |
| ≥12 | 1083 (11·3) | 65·0 (61·1-65·0) | 92·4 (91·9-93·0) | 0·4 | 8·6 | 37·1 | 97·5 | 37·1 |
| ≥13 | 604 (6·3) | 46·3 (42·3-50·3) | 96·5 (96·1-96·8) | 0·6 | 13·1 | 47·4 | 96·3 | 47·4 |
| ≥14 | 276 (2·9) | 28·2 (24·6-31·9) | 98·9 (98·6-99·1) | 0·7 | 24·8 | 63·0 | 95·2 | 63·0 |
| ≥15 | 100 (1·0) | 13·1 (10·5-16·0) | 99·8 (99·7-99·9) | 0·9 | 62·0 | 81·0 | 94·4 | 81·0 |
| ≥16 | 29 (0·3) | 4·4 (2·9-6·3) | 100 (99·9-100) | 1·0 | 196·3 | 93·1 | 93·8 | 93·1 |
| ≥17 | 5 (0·1) | 0·8 (0·3-1·9) | 100 (100-100) | 1·0 | - | 100·0 | 93·6 | 100·0 |

FN=false negative, FP=false positive, NPV=negative predictive value, PPV=positive predictive value, TN=true negative, TP=true positive.

**Appendix Table 5.**
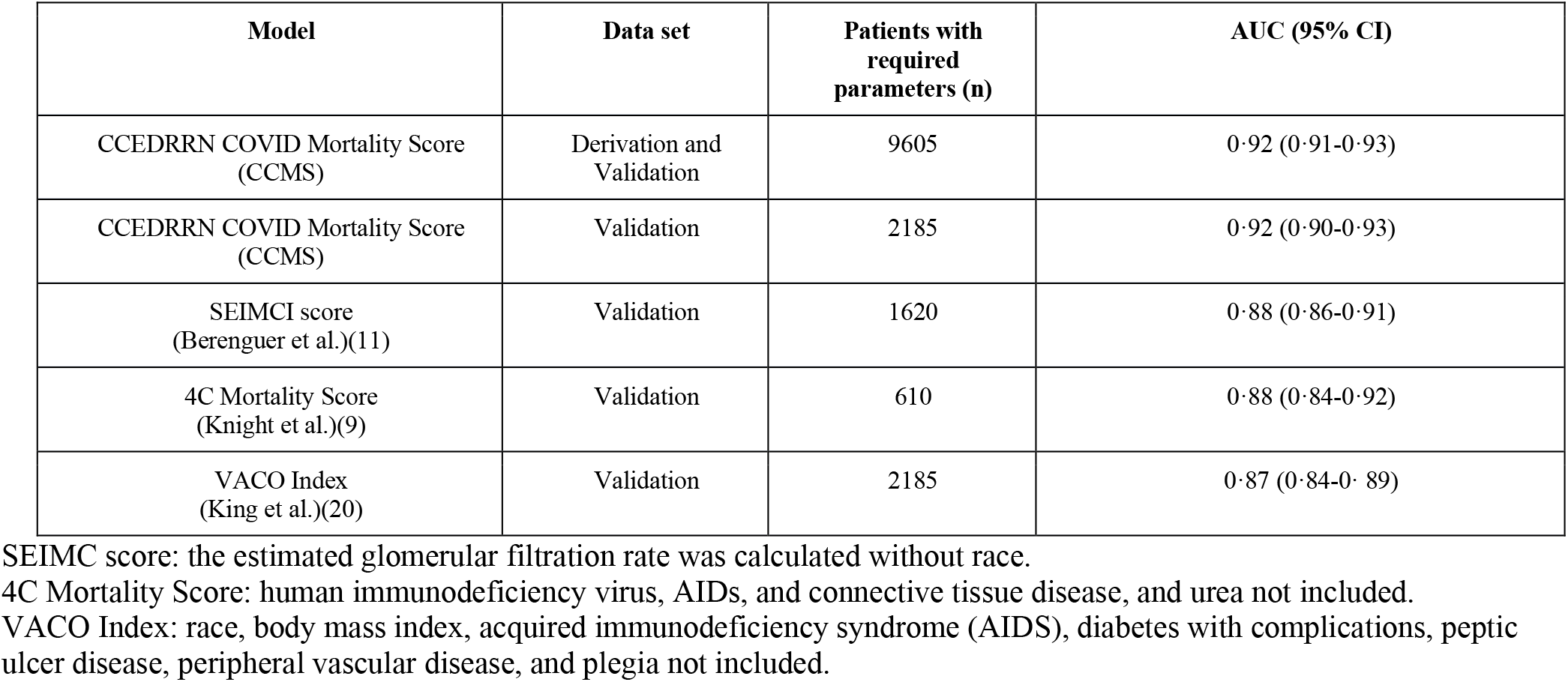
Receiver operator curves and area under the curve (AUC) for the CCEDRRN COVID Mortality Score, the SEIMC score, the 4C Mortality Score, and the VACO Index.

| Model | Data set | Patients with required parameters (n) | AUC (95% CI) |
| --- | --- | --- | --- |
| CCEDRRN COVID Mortality Score (CCMS) | Derivation and Validation | 9605 | 0·92 (0·91-0·93) |
| CCEDRRN COVID Mortality Score (CCMS) | Validation | 2185 | 0·92 (0·90-0·93) |
| SEIMCI score (Berenguer et al.)(11) | Validation | 1620 | 0·88 (0·86-0·91) |
| 4C Mortality Score (Knight et al.)(9) | Validation | 610 | 0·88 (0·84-0·92) |
| VACO Index (King et al.)(20) | Validation | 2185 | 0·87 (0·84-0·89) |

SEIMC score: the estimated glomerular filtration rate was calculated without race.
4C Mortality Score: human immunodeficiency virus, AIDs, and connective tissue disease, and urea not included.
VACO Index: race, body mass index, acquired immunodeficiency syndrome (AIDS), diabetes with complications, peptic ulcer disease, peripheral vascular disease, and plegia not included.

**Appendix Figure 1·.**
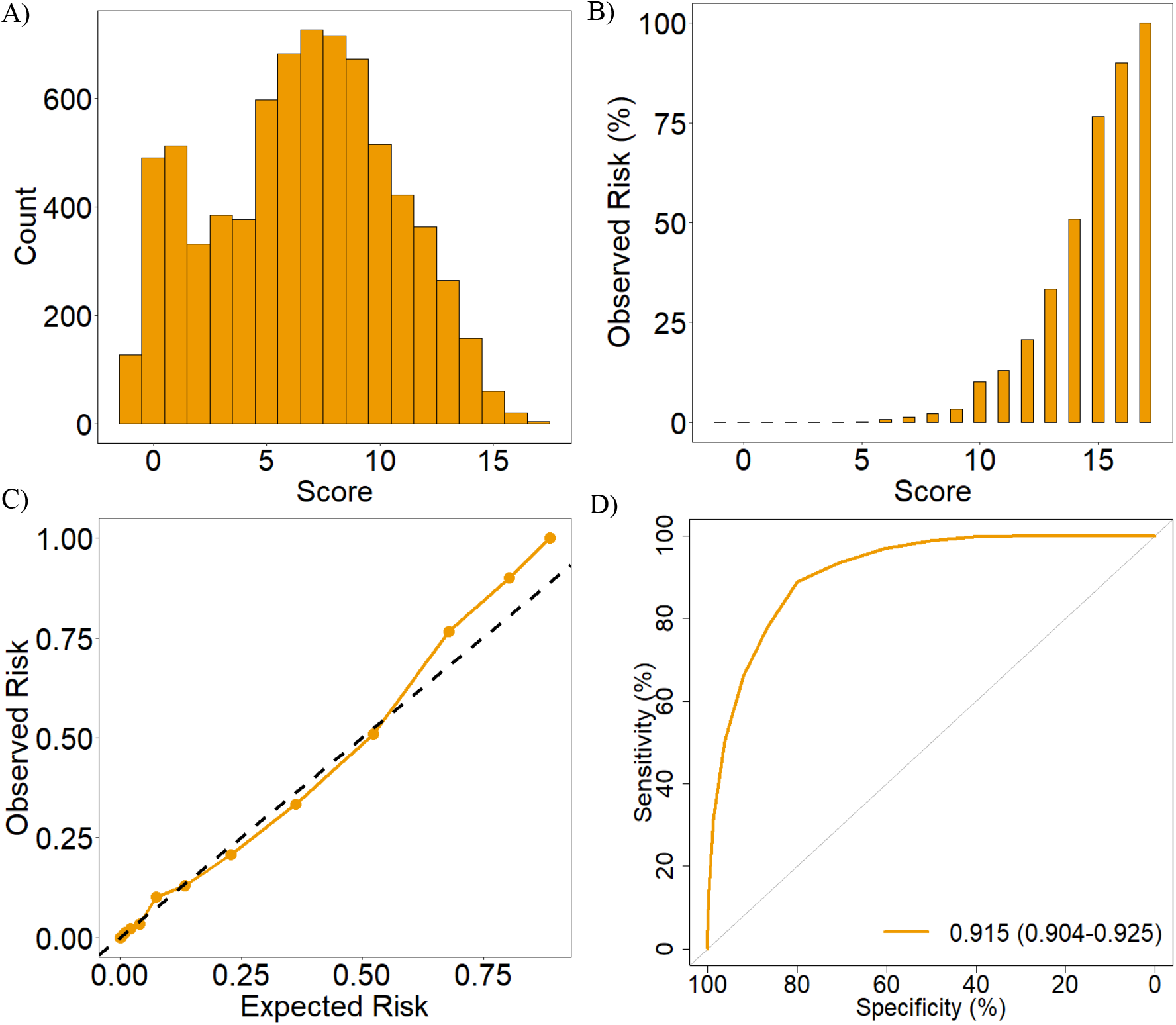
Distribution and performance of the CCEDRRN COVID Mortality Score in the derivation cohort: A) distribution of the score, B) observed in-hospital mortality across the range of the score, C) predicted versus observed probability of in-hospital mortality, and D) receiver operating characteristic curve with area under the curve (AUC) and associated 95% confidence interval.

